# Patient perspectives on living with hypertension: Social media listening analysis across predominantly high-income countries

**DOI:** 10.64898/2026.04.22.26351483

**Authors:** Salvatore Di Somma, Rachel Gervais, Marc Bains, ShantaQuilette Carter-Williams, Simon Messner, Naomi Onsongo

## Abstract

**Background:** Chronic conditions such as hypertension can significantly disrupt daily life and emotional well-being. The interaction between patients’ perceptions, adherence to antihypertensive medication and quality of life (QoL) remains underexplored outside structured clinical settings.

**Objectives:** To capture unprompted patient perspectives and assess whether hypertension affects QoL and to investigate if patient reported experiences are associated with self-reported antihypertensive medication adherence.

**Methods:** Social media listening (SML) study analyzing 86,368 anonymized posts from individuals with hypertension in 12 countries, collected between January 2022 and May 2024. Posts from 11 countries (n=81,368) were analyzed using artificial intelligence–enabled natural language processing. Posts from China (n=5,000) were analyzed separately using a harmonized framework. Quantitative and qualitative methods assessed variations by country, age, and gender, and associations between emotional expression and antihypertensive medication adherence.

**Results:** Across the 11-country core sample, 45% of posts mentioned at least one QoL impact, most commonly worry/anxiety (11%). Impacts varied across countries. Among 8,096 posts with age identified, individuals <40 years reported emotional balance impacts in 28% of posts versus 22% among those aged 40+. Work/Education impacts were mentioned in 17% of posts by those <40 years vs 12% in 40+. Among 7968 posts explicitly referencing adherence, expressed worry was associated with stricter adherence (62% association score), as were structured routines (79% score), home monitoring (77%), dietary changes (77%), and exercise (71%). In contrast, sadness/depression was associated with inconsistent adherence (71%), as were forgetfulness (79%), side effects (73%), and cost/insurance concerns (65%).

**Conclusions:** These results emphasize the importance of the psychological and emotional impact of hypertension, including on adherence to medication regimens, reinforcing the value of a holistic approach to patient care.

**Plain language summary:** Many people have high blood pressure, which increases the risk for stroke and other harmful events. Although there is much medical research on high blood pressure, little is known about the experiences of those people who live with the condition. This study aimed to scope out how people react emotionally and how their high blood pressure affects their daily lives. To do this, we analyzed social media posts from the United States, Canada, Brazil, the United Kingdom, Germany, France, Italy, Spain, Japan, South Korea, China, and Australia. In all countries, patients were affected emotionally by their high blood pressure. People often worried, particularly when they received their diagnosis. High blood pressure also negatively affected everyday life and work/education. This was often due to frequent medical appointments with lengthy wait times and difficulty accessing specialists. Taking medications regularly and sticking to diet and exercise programs was more difficult for individuals who reported being sad or depressed, or who suffered from side effects of their medications. The findings show that efficient management of high blood pressure has to take into account the emotional reactions of those who are affected, and provide support in several areas beyond the prescription of medication.

## Introduction

Often described as a “silent killer” due to its frequent lack of noticeable symptoms, hypertension is the leading risk factor for death and disability globally. ^1,2^ Hypertension affects approximately one in three people worldwide,^2^ and is expected to become increasingly prevalent as global populations age and certain risk factors, such as obesity, become more widespread.^1^ Hypertension remains poorly controlled, with non-adherence to medication or lifestyle interventions identified as a significant barrier to effective disease management.^3,4^ An understanding of the patient experience is fundamental for the provision of individualized interventions and measures to support adherence. Although traditional research methods, including qualitative interviews, quantitative surveys and data from clinical research, have provided invaluable insights,^3,5^ these approaches may fail to fully capture patients’ subjective experiences and the broader impacts of the condition.

Social Media Listening (SML), the systematic analysis of unsolicited online conversations, is a powerful noninterventional research technique, enabling the study of authentic, unprompted, organic patient narratives at scale.^6–8^ Recognized by regulatory bodies such as the US Food and Drug Administration (FDA) as a potential source of patient experience data,^9^ SML has been employed to assess quality of life (QoL) impacts,^8,10^ and explore antihypertensive medication adherence across geographically and demographically diverse populations.^6,10^ However, to date, no published SML studies have specifically investigated patient perspectives on hypertension experiences or management, highlighting a critical gap in current knowledge. The current study employs SML and advanced Natural Language Processing (NLP) techniques to analyze patient perspectives on living with hypertension across twelve countries.

## Methods

This was a cross-sectional SML with the primary objective to characterize whether hypertension affects patients’ QoL and emotional well-being, and to examine if these reported experiences are associated with self-reported adherence to antihypertensive medication. A secondary aim was to explore variations across demographic groups and country contexts.

### Data Collection and preparation

SML was used to capture publicly available posts from individuals with hypertension in the United States (US), Canada, Brazil, the United Kingdom (UK), Germany, France, Italy, Spain, Japan, South Korea, China, and Australia between January 2022 and May 2024). Posts were collected from global platforms (e.g., Facebook, Reddit, X, Instagram), Chinese platforms (Xiaohongshu, Weibo, Douyin, Bilibili), and country-specific patient forums. In China, WeChat was not among the sources, as much of the activity on this platform takes place within private environments and are not accessible for data collection due to privacy restrictions.

Posts were identified using an English-language keyword query developed by the analyst team for hypertension-related patient discussions, translated into eight languages (Portuguese, German, French, Italian, Spanish, Japanese, Korean, Mandarin Chinese). Eligibility criteria required posts to be authored by individuals with hypertension, publicly available, and published within the study timeframe. eligible posts meeting these criteria within the timeframe were included. Posts were excluded if they were duplicates (e.g., cross-posts, retweets), metaphorical (e.g., non-clinical uses of “high blood pressure”), or lacking sufficient patient-experience detail. Posts published in languages other than English were systematically translated using Google Translate (version 2.0.14) and then reviewed and refined by multilingual analysts to ensure accurate rendering and semantic faithfulness.

To approximate relative hypertension prevalence and national patient population sizes, the dataset was rebalanced by proportionally removing excess posts (∼3,000 in total) from countries with disproportionately high digital verbosity. To enable comparisons between countries and populations (e.g., gender differences), all posts from the two-year period were included.

Data anonymization complied strictly with the General Data Protection Regulation (GDPR) principles, involving automated removal of social media handles and manually checking for potentially identifying information (names, specific locations, or distinctive personal details). All analyses were performed on fully anonymized datasets.

As this study utilized existing, publicly accessible, and anonymized data, the independent ethics committee Pearl IRB (Indianapolis, USA) determined that it did not constitute human subjects research (ID 2024-0294) and was thus deemed exempt of IRB approval.

### Language data analysis

Analysis utilized Luminoso, (Luminoso Technologies INC, Boston USA)^11^ a commercially available artificial intelligence-driven NLP platform. Data from China were analyzed separately, following research frameworks and themes developed in the core analysis, to comply with China data transfer regulations. Luminoso was selected for its capacity to robustly explore semantic nuance in large-scale text data sets through inductive (bottom-up) thematic analysis.^12^ Luminoso surfaced clusters of semantically related words and phrases based on co-occurrence patterns. The research team manually reviewed these outputs to define “concepts” (e.g., “anxiety”) and iteratively grouped them into broader thematic areas (e.g., “emotional impacts”) and analytical domains (e.g., “QoL impacts”) following standard patient-experience research frameworks.^8^ All identified concepts and groupings were independently reviewed by analysts not involved in their creation. Discrepancies were resolved by consensus. In total, approximately 130 individual concepts (grouped into the analytical domains of disease terms and status comorbidities, broader perceived causes, symptoms, life impacts, non-RX disease management, medications, side effects, healthcare professional (HCP) types, and journey stages) were analyzed. Antihypertensive adherence was defined as consistent daily use of prescribed medication, with inconsistent adherence identified as any deviation from this routine (eg missed doses).

Demographic variables were coded where identifiable. For age, posts were categorized as <40 years or 40+ years, in agreement with the BMJ definition of young-onset hypertension as <40 years^13^ and American Heart Association age groupings.^14^

### Statistical analysis

The final dataset was examined using mixed-methods analysis combining quantitative metrics with qualitative review of patient language and illustrative quotations, following best practices in SML research.^8^ Association scores to quantify the probability that posts mentioning one concept also mention a second concept, were calculated using concept thresholds defined as negligible (<2%), uncommon (2–6%), quite common (7–20%), common (21–40%), and very common (>40%.

Primary outcomes were the frequency and thematic patterns of QoL impacts, emotional expressions, and antihypertensive medication adherence. Predictors included age group, gender, and country where identifiable. No adjustment for confounders was possible due to limited demographic detail in social media posts.

## Results

### Sample size and demographics

A final sample of 86,368 social media posts from 67,200 unique accounts were analyzed. The core sample used for Luminoso language analysis was 81,368 posts, comprising content from 11 countries (excluding China) and an additional 3,700 English-language posts on Reddit that mentioned primary hypertension or hypertension with renal comorbidities (Table 1). An additional 5,000 posts from China were collected and analyzed separately.

**Table 1.** Distribution of posts included in the SML study.

| Variables and categories | Posts, n (%) |
| --- | --- |
| **Country** | **n=86,368** |
| US | 36,307 (42%) |
| Brazil | 11,209 (13%) |
| Canada | 2,430 (3%) |
| UK | 4,589 (5%) |
| Germany | 4,194 (5%) |
| France | 2,686 (3%) |
| Italy | 1,286 (1%) |
| Spain | 5,059 (6%) |
| Japan | 5,486 (6%) |
| South Korea | 3,517 (4%) |
| China | 5,000 (6%) |
| Australia | 915 (1%) |
| Unclassified by location Reddit boost (English-language) | 3,690 (4%) |
| **Gender** | **n=22,394** |
| Female | 11,519 (51%) |
| Male | 10,875 (49%) |
| **Age** | **n=8096** |
| Under 40 Years Old | 2,716 (34%) |
| 40 Years Old and Above | 5,380 (66%) |
| **Ethnicity** | **n=794** |
| White | 269 (34%) |
| Black | 240 (30%) |
| Asian | 153 (19%) |
| Hispanic | 132 (17%) |

The core sample consisted of similar proportions of male (51%) and female gender. Two-thirds 66%) were aged 40+ years (Table 1). Ethnicity information was available for <1% of the posts. In all reported results, unless explicitly stated, values reflect the 11 countries core sample excluding China.

### Terminology used by patients to describe their condition

Across the core sample, patient posts used the lay term “high blood pressure” (86%) rather than the clinical term “hypertension” (14%) when describing their condition. The clinical term “hypertension” was more commonly used by patients with severe conditions or comorbidities, who explicitly mentioned consultations with cardiologists, potentially reflecting their increased exposure to medical terminology. Patients explicitly referred to a diagnosis of “primary” or “essential” hypertension in 10% of posts. The term “secondary” hypertension was rarely mentioned.

### Condition status and comorbidities

Of the 13,005 posts mentioning hypertension control, 88% referred to the condition as controlled by treatment or management. Cardiovascular conditions and heart problems were most common comorbidities mentioned, occurring in 24% of all core sample posts. Metabolic conditions (19%) were also commonly mentioned; of these, diabetes (14%) and obesity (8%) were most common.

> “Probably linked to my obesity, I have been diagnosed with hypertension (180/90 without drugs) as well as prediabetes” - Patient, Primary hypertension, Male, 40+ age group, United States.

Psychiatric conditions (9%) were commonly mentioned, with anxiety disorders (6%) and depression (3%) most common (details in Supplementary materials).

> “I have been struggling with anxiety, panic disorder and major depressive disorder for 14 years now. Over the years I have also developed hypertension.” - Patient, Hypertension with cardiac comorbidities, Female, <40 age group, United Kingdom.

### Symptom mentions

Hypertension-related symptoms were rare in the sample. A long tail of 15 distinct physical symptoms was identified, each mentioned in no more than 5% of posts. Most common individual symptoms attributed directly to hypertension (rather than to medication side effects) were joint or muscle pain (5%), headaches or migraines (4%), hearing issues (4%) including tinnitus and other sounds and vision disturbances (3%) (Supplementary materials). Patients who mentioned their hypertension status (controlled, uncontrolled, or resistant) also discussed symptoms more frequently. Those with uncontrolled or resistant hypertension were more likely to report symptoms, particularly aches and pain (16% vs. 10%) and fatigue and sleep issues (11% vs. 5%) compared to those with controlled hypertension (details in Supplementary materials).

### Quality of Life impact mentions

QoL impacts were mentioned frequently (in 45% of all core sample posts) spanning the areas of everyday life (21%), emotional balance (20%), work/education (10%), social connections (6%), and financial health (5%) (Table 2). Patient posts frequently contained references to more than one impact area. The single most common individual impact was worry/anxiety (11%), which was mentioned especially frequently at diagnosis (15% of all worry/anxiety mentions).

**Table 2.** Quality-of-life impacts in 81,368 social media posts on hypertension from 11 countries.

|  |  | Percent % of posts |
| --- | --- | --- |
| Life Impact Mentions |  |  |
| **Total Life Impacts (Any Mention)** |  | 45 |
| **Life Impact Area:** | **Everyday Life (total)** | 21 |
| Individual Life | Time spent on medical appointments | 9 |
| Impacts: | Sleep disruption | 6 |
|  | Time spent researching online | 5 |
|  | Functional limitations | 3 |
|  | Household task difficulties | 2 |
| **Life Impact Area:** | **Emotional Balance (total)** | 20 |
| Individual Life | Worry and anxiety | 11 |
| Impacts: | Sadness and depression | 4 |
|  | Fear | 2 |
|  | Shock and disbelief | 2 |
| **Life Impact Area:** | **Work/ Education (total)** | 10 |
| Individual Life |  |  |
| Impacts: | Impact on job performance | 9 |
| **Life Impact Area:** | **Social Connections (total)** | 6 |
| Individual Life | Impact on family & friendships | 3 |
| Impacts: | Ability to maintain a social life | 2 |
| **Life Impact Area:** | **Financial Health (total)** | 5 |
| Individual Life | Expense cost concerns | 3 |
| Impacts: | Insurance coverage | 2 |

Although the differences were small, Patients in Spain, France, Germany, South Korea and Japan reported impacts with higher frequency than the global average (Table 3). In Europe, patients from Spain, France, and Germany frequently highlighted impacts related to everyday life, emphasizing practical challenges such as the inconvenience, time burden, and disruption to leisure or personal activities caused by frequent medical appointments with lengthy wait times and difficulties accessing specialists. Financial concerns were mentioned in <10% of posts across countries.

**Table 3.** Cross-country variation in QoL impacts in 81,368 social media posts from all 12 countries. Multiple mentions are possible.

| Country | Total QoL<br>Impacts n<br>(%) | Everyday<br>Life (%) | Emotional<br>Balance<br>(%) | Work &<br>Education<br>(%) | Social<br>Connections<br>(%) | Finances<br>(%) |
| --- | --- | --- | --- | --- | --- | --- |
| Global Sample | 45 | 21 | 20 | 10 | 6 | 5 |
| South Korea n= 3,517 | 64 | 28 | 29 | 18 | 10 | 8 |
| France n= 2,686 | 60 | 33 | 25 | 11 | 11 | 6 |
| Spain n= 5,059 | 55 | 34 | 17 | 11 | 10 | 6 |
| Germany n= 4,194 | 51 | 25 | 27 | 11 | 7 | 3 |
| Japan n= 5,486 | 48 | 20 | 21 | 10 | 7 | 7 |
| United Kingdom n= 4,589 | 45 | 21 | 19 | 10 | 5 | 4 |
| Canada n= 2,430 | 45 | 21 | 19 | 13 | 6 | 3 |
| Australia n= 915 | 44 | 20 | 21 | 11 | 4 | 6 |
| United States n= 36,307 | 42 | 19 | 19 | 10 | 5 | 5 |
| China n= 5,000 | 41 | 10 | 15 | 10 | 7 | 7 |
| Brazil n= 11,209 | 38 | 12 | 21 | 5 | 5 | 4 |
| Italy n= 1,286 | 31 | 20 | 17 | 5 | 7 | 4 |

### Subgroups

Within the 8096 core sample posts that could be categorized by age, patients self-identifying as below 40 years of age more frequently mentioned cardiovascular issues and diabetes as co-conditions, with obesity more common than in those aged 40+ years (details in Supplementary materials). Worry/anxiety, which was the most commonly mentioned impact, was mentioned more frequently by patients aged below 40 years (19% vs. 13% overall), and this group also more frequently reported impacts on their emotional balance (28% vs. 22%), work/ education (17% vs. 12%), and social connections (9% vs. 5%) than their older counterparts (Table 4). Whilst many patients aged 40+ years tended to describe hypertension in resigned terms, it was common for younger patients to express high levels of concern about the implications of their diagnosis for their broader health and lifestyle.

**Table 4.**
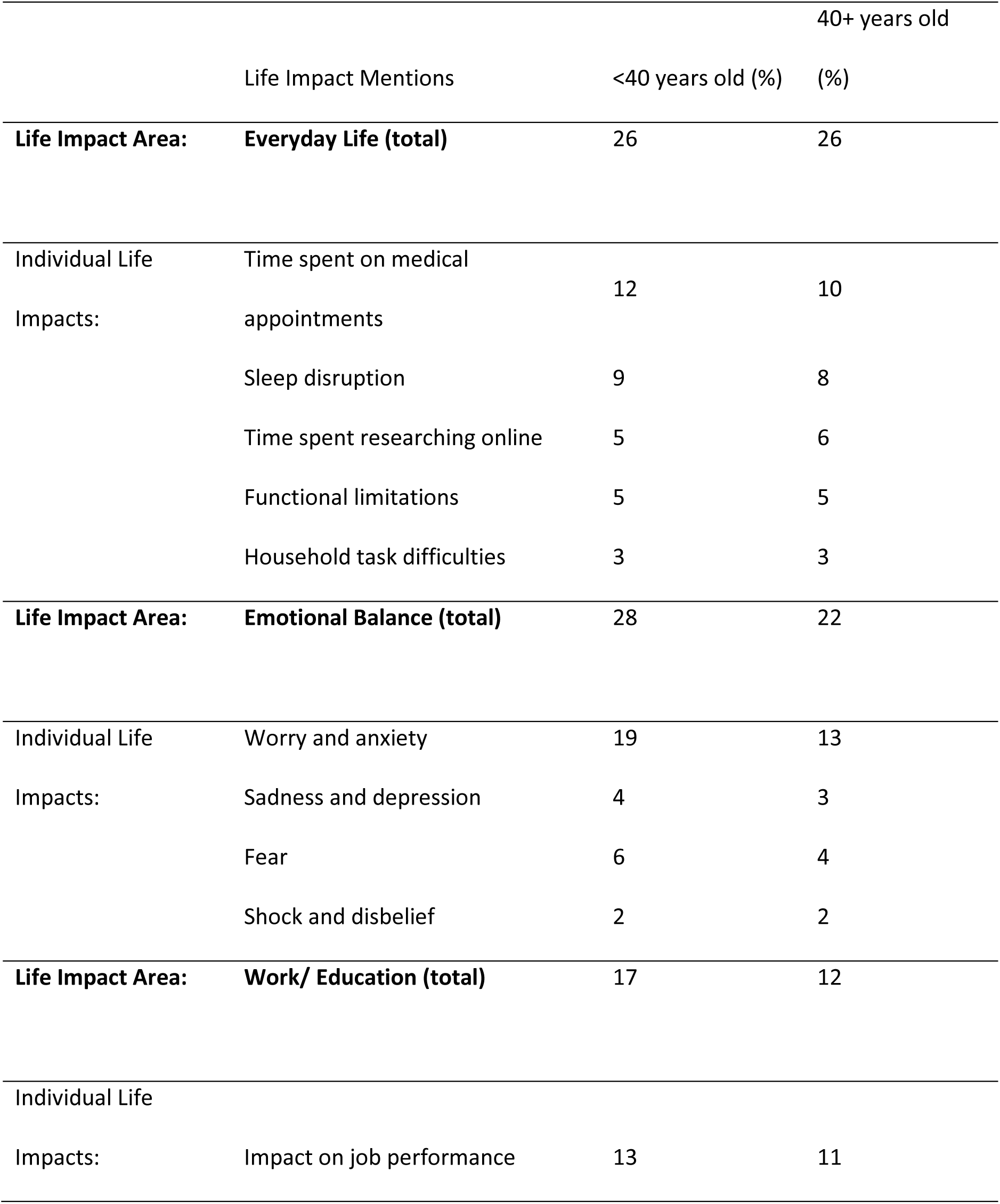
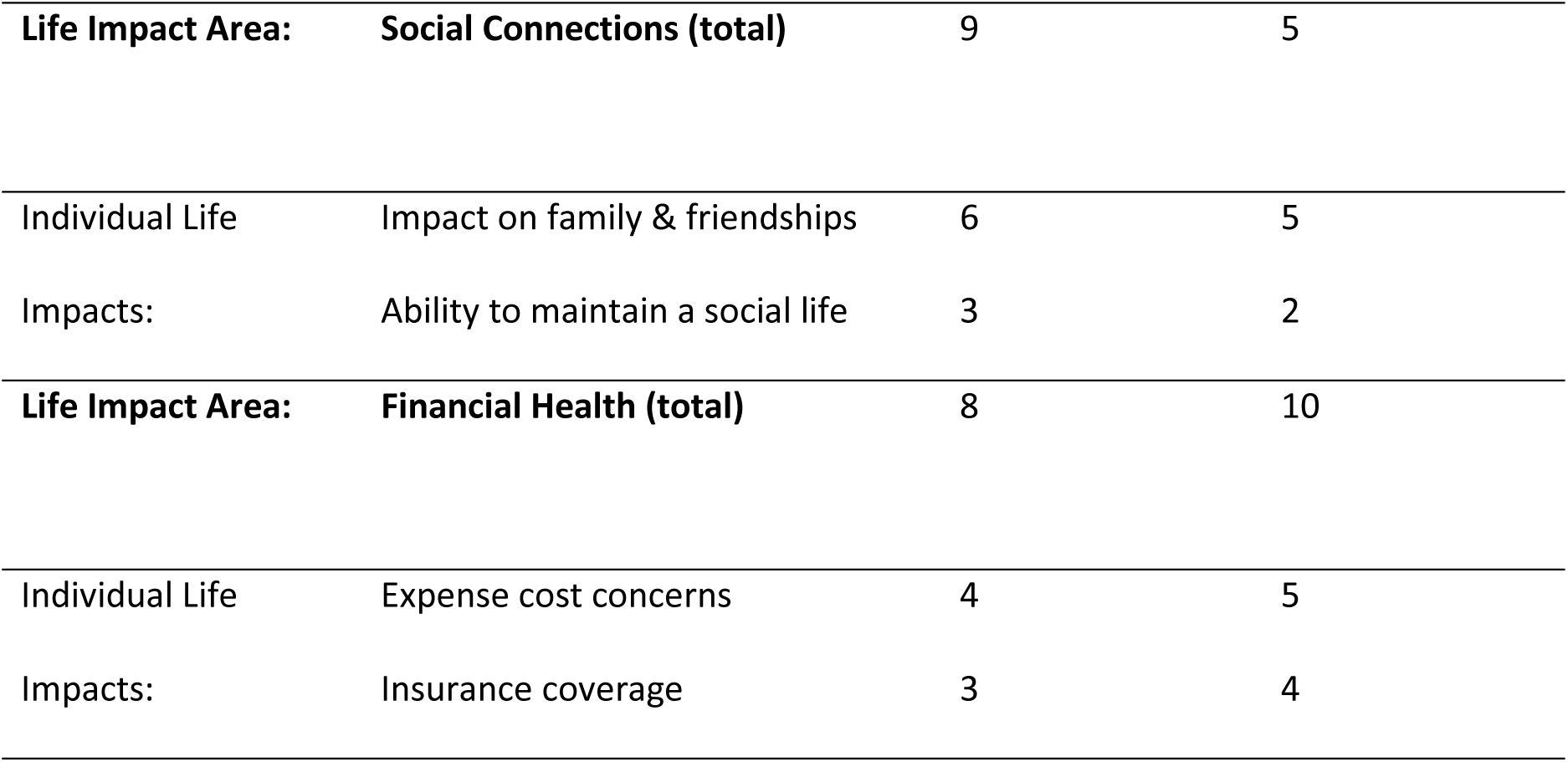
Age-group variation in QoL impacts in 8096 patient social media posts on hypertension from 11 countries excluding China, January 2022–May 2024. Percentages reflect posts per age group mentioning at least one life impact area.

> *“When they told me I had HBP I said nope, I’m too young for this, this will drastically change my life”* - Patient, Male, <40 years age group, Canada.

Additionally, patients below 40 years of age more frequently mentioned individual impacts such as negative effects of hypertension on their job performance (13% vs 11%) and the disruption of medical appointments (12% vs 10%) to their daily life than older patients. Patients aged 40+ years more frequently highlighted financial health concerns related to medical expenses, insurance coverage and financial security in retirement (Table 4).

> *“After retiring at 60 I had to face reduced healthcare access and higher costs of treatment”* - Patient, Hypertension with cardiac comorbidities, Male, 40+ age group, China.

Patients with concomitant cardiovascular conditions more frequently cited everyday life impacts on QoL (mentioned in 28% of posts), notably time burdens from medical appointments and online research, as well as activity limitations compared to the general hypertensive population (details in Supplementary materials).

Across the 22,394 core sample posts that could be categorized by gender, differences in the frequency of reported QoL impacts were minor (details in Supplementary materials).

Very few posts (n=794; all in the US) included self-identified race or ethnicity. There appeared to be signs of lower quality of care provided to Black patients compared with White and Hispanic patients, but the dataset was too limited for detailed analysis (details in Supplementary materials).

### Antihypertensive medication adherence

Among the 7968 core sample posts (9% of total) explicitly referencing antihypertensive medication adherence, 54% described strict medication adherence, whereas 46% indicated inconsistent medication adherence, such as through mentions of missed doses.

More adherent patients emphasized consistent routines and frequently highlighted structured monitoring methods.

> *“I focus on taking my meds and going for daily walks – I try not to let the good habits slip”* – Patient, Hypertension with cardiac comorbidities, Unknown gender, 40+ age group, France.

> *“I have an app on my phone. Just to keep an eye on it.”* – Patient, Primary hypertension, Unknown gender, <40 age group, Japan.

Reported medication adherence was associated with a range of proactive self-management behaviors (Table 5) such as structured daily routines and habits, at-home monitoring of the condition, dietary changes, and physical exercise. Posts also frequently mentioned proactive online research and regular contact with HCPs.

**Table 5.** Associations between mentions of antihypertensive medication adherence and related behaviors in 7968 patient social media posts on hypertension from 11 countries (excluding China).

| Adherence Drivers | Association Score (%) |
| --- | --- |
| **Consistent Antihypertensive Medication Adherence** |  |
| Structured daily routines and habits | 79 |
| At-home condition monitoring | 77 |
| Dietary changes | 77 |
| Physical exercise | 71 |
| Independent online research | 69 |
| Worry/ anxiety | 62 |
| Severe condition | 54 |
| **Less Consistent Antihypertensive Medication Adherence** |  |
| Forgetfulness | 79 |
| Medication side effects | 73 |
| Sadness / depression | 71 |
| Cost/ insurance issues | 65 |
| Milder condition | 65 |

Emotional concern was more strongly tied to adherence than disease severity: mentions of worry and anxiety were frequently co-mentioned with strict adherence (62% association) compared with a 54% association between mentions of severe or complex conditions and adherence. The emotional impact of sadness or depression was associated with less consistent antihypertensive medication adherence (71% association score).

The analysis highlighted several key barriers to antihypertensive medication adherence (Table 5). In addition to general forgetfulness (79% association score) and the perception of having a mild condition (65% score), patients commonly mentioned medication side effects (73% association) and cost or insurance-related issues (65%).

> *“I have been on blood pressure medication for years but often forgot to take it or run out. Even when I ran out of pills before, I didn’t go to the hospital”* – Patient, Hypertension with renal comorbidities, Unknown age & gender, Japan.

> “*I have been prescribed blood pressure meds but I’m going to experiment to see if I actually need them. I have made some changes to my diet.”* - Patient, Hypertension with cardiac comorbidities, Unknown age & gender, United Kingdom.

> *“Then I experienced the tablet side effects, like impotence… dizziness, fatigue. I stopped taking my tablets on my own, without consulting the doctor.”* - Patient, Unknown gender, <40 age group, Germany.

## Discussion

This cross-sectional SML study found that hypertension has substantial impacts on patients’ QoL. Despite its often hidden clinical profile, hypertension was found to carry a high emotional toll similar to other chronic illnesses, even those with minimal or no overt symptoms.^15–18^ Within this dataset, hypertension was linked to psychological and practical burdens even without comorbidities. Such findings challenge assumptions that QoL are primarily comorbidity-driven and reinforce the need to view hypertension itself as a condition with significant standalone impact on patients’ lives. To our knowledge, this is the first large-scale, multinational SML study which systematically investigates patient perspectives on hypertension experiences and management.

The adherence rates aligned with the 45.2% adherence rates reported in a global meta-analysis of 25 research studies.^19^ Worry and anxiety were associated with strict adherence, but depressive symptoms were associated with less consistent adherence. This second finding complements a meta analysis in 2011 which estimated the odds of a depressed patient being non-adherent to 1.76 times the odds of a non-depressed patient.^20^ Studies have shown that when patients perceive a condition as asymptomatic or low-risk, as is often the case with hypertension, they may struggle to maintain adherence due to a lack of urgency.^21–23^ Clearly, the relationship between emotional impact and behavioral patterns is complex and needs further studies to untangle. Differences were also observed across countries, age groups, and genders, all of which would seem relevant to explore in further studies.

Patients below 40 years of age more frequently reported significant emotional impacts, including anxiety, worry, and social disruption, than older patients. This finding aligns with broader evidence from other chronic conditions, which suggests that younger individuals typically experience greater distress, partly due to the unexpected nature of chronic illness early in life.^24^ The differences in specific impact were too small to draw strong conclusions, and would need further research to guide age-adapted disease-management strategies that address both emotional and lifestyle-related impacts.

There is a clear need for a nuanced approach to support emotional engagement in hypertensive patients without inducing counterproductive stress responses, which have been shown to worsen blood pressure outcomes.^25,26^ An emotional impact assessment is desirable for hypertension management and for the design of interventions and communications that are responsive to local health system contexts and patient expectations, enabling more relevant and equitable hypertension care. Patient associations could play a valuable role, raising awareness of patient needs, tackling potential stigma around emotional disease impacts, and providing education and support to patients to improve their QoL. The dual role of emotion as both a motivator and a potential barrier underscores the complexity of addressing psychological factors in chronic disease care. Whilst patients may be motivated by concern about their condition and report stricter condition management practices, they may also be affected by competing emotional pressures such as frustration with side effects or anxiety over cost and insurance coverage. Prior research has similarly identified economic strain and adverse treatment experiences as major contributors to non-adherence.^27,28^ These counter concerns may blunt the motivational effect of worry, leading some patients to be less consistent in their condition management.

Quantitative data from SML differ significantly from traditional survey data, as social media posts typically reflect patients’ most immediate or significant concerns rather than exhaustive experiences. Thus, the frequencies of mention in social media data likely represent more common experiences than equivalent frequencies from structured surveys. Drawing upon previous experience with SML studies,^29^ the research team assumed that experiences mentioned in approximately 50% of posts might indicate near-universal patient prevalence. This is not a hard threshold however: mention frequencies varied substantially by topic; for example, symptoms were generally discussed more frequently than less immediately salient topics, such as comorbidities or medications, which may reflect their experienced importance rather than actual prevalence.

### Strengths and limitations

While SML provides valuable insights into authentic patient experience, several limitations warrant consideration. Demographics within each country’s sample were intentionally not adjusted to preserve an accurate representation of individuals publicly discussing hypertension online. Self-reported content may mis-state demographics or clinical detail. Selection bias is likely: public posters tend to be younger, more digitally engaged, and more willing to discuss health, potentially underrepresenting older adults and those less inclined to share, which can affect representativeness.^10^ Among other potential sources of bias are platform bias (posting norms and moderation shape what is visible), keyword/translation bias (query terms and multilingual renderings preferentially surface some topics/voices), and algorithmic bias (machine translation and NLP clustering may misclassify or overweight frequent terms). We mitigated these risks through multi-platform capture, multilingual analyst review, proportional rebalancing across countries, and independent analyst consensus checks, but residual bias is likely.^6,7,10^

Associations between variables do not necessarily imply causation and as the study design did not include control groups or follow-up, comparisons should be made with care. For similar design reasons, the statistical significance of associations could not be calculated. As noted above, comparisons between frequencies observed with an SML methodology and traditional surveys are fraught.

Among strengths are the huge number of data points and the ability of SML to capture extensive, unmediated patient perspectives at scale,^6,10^ which is a valuable complement to traditional patient-experience methodologies. Similarly, given hypertension’s often asymptomatic nature and the potential stigma associated with antihypertensive medication non-adherence (e.g. the fear of being judged as non-compliant),^4^ patients may underreport their true experiences or adherence challenges in clinical settings or structured research context but be more open about such issues in peer-to-peer discussions.

Geographic scope was a major constraint: the study sample was drawn primarily from high-income countries, excluding regions most severely affected by the global hypertension crisis such as Southeast Asia and Africa. The findings should not be uncritically generalized to low- and middle-income countries. Future research should expand representation to these regions to capture cultural and socioeconomic variation in experience and adherence.

## Conclusions

The findings in this study emphasize the necessity of explicitly acknowledging and addressing the psychological and emotional impacts of hypertension which is traditionally considered asymptomatic, and reinforce the value of a holistic approach to patient care. Notably, within this cross-sectional SML dataset, the expression of emotional concern was associated with self-reported adherence patterns.

Further research should build on these findings and examine the interplay between patient perceptions, emotional experiences, and medication adherence. An understanding of how to balance motivating concern with avoiding excessive worry or anxiety that can induce harmful stress and worsen outcomes will be crucial for advancing comprehensive patient care and adherence strategies.^25,26^

## Acknowledgements

The authors thank members of the Novartis patient insights panel, Flavia Perna, Ramandeep Jindal, Andrey Shutov, and Anne Olshan for their review and ideation on publication planning.

The authors acknowledge the use of ChatGPT, accessed in May 2025, in assisting with the refinement of the English in the paper. After using this tool, the authors reviewed and edited the content as necessary and assume full responsibility for the paper’s content. This study and publication were funded by Novartis Pharmaceuticals Corporation.

## Author contributions

All authors made a significant contribution to the work reported, whether that is in the conception, study design, execution, acquisition of data, analysis and interpretation, or in all these areas; took part in drafting, revising or critically reviewing the article; gave final approval of the version to be published; have agreed on the journal to which the article has been submitted; and agree to be accountable for all aspects of the work.

## Data availability

The datasets generated during and analyzed during the current study are not publicly available due to legal and privacy considerations.

## Abbreviations

HCP: Healthcare Professional
SML: Social Media Listening
QoL: Quality of Life

