## Supplementary materials for "Patient perspectives on living with hypertension: Social media listening analysis across predominantly high-income countries"

**English Keyword Query**

(
(
// BASIC HYPERTENSION TERMS //
(“high blood pressure” OR #highbloodpressure OR “increased BP” OR “increased blood pressure” OR HBP OR #HBP OR “elevated blood pressure” OR “elevated BP” OR “my BP”)
OR
(// ESSENTIAL HYPERTENSION //
(“primary hypertension” OR primaryhypertension OR #primaryhypertension OR #primary #hypertension OR “primary high blood pressure” OR “essential hypertension” OR essentialhypertension OR #essentialhypertension OR “familial hypertension” OR “familial high blood pressure” OR “genetic hypertension” OR “genetic high blood pressure” OR “hereditary hypertension” OR “hereditary high blood pressure” OR “inherited hypertension” OR “inherited high blood pressure” OR “predisposed to hypertension” OR “predisposed to high blood pressure”)
OR
(
(
(“high blood pressure” OR #highbloodpressure OR hypertension OR #hypertension OR “hypertensive crisis” OR #hypertensivecrisis OR “hypertensive emergency” OR #hypertensiveemergency OR #bloodpressure OR #highbloodpressure OR #hypertensiontreatment OR “antihypertensive drugs” OR antihypertensive OR #antihypertensive OR “DASH diet” OR #DASHdiet OR HBP OR #HBP OR #hypertensionawareness)
AND
(// IDIOPATHIC //
idiopathic OR #idiopathic OR idiopathy OR #idiopathy OR “unknown cause” OR “no known cause” OR “don't know the cause” OR “dont know the cause” OR “don't know cause” OR “dont know cause” OR
// FAMILIAL //
genetic OR familial OR #genetic OR #familial OR “runs in my family” OR “runs in their family” OR “ran in my family” OR “ran in their family” OR “genetic predisposition” OR “genetically predisposed” OR gene OR “I inherited” OR hereditary OR “inherited from” OR
// CONFOUNDING FACTORS //
obesity OR #obesity OR obese OR #obese OR overweight OR #overweight OR “lack of exercise” OR “never exercise” OR “dont exercise” OR “don't exercise” OR “I am not fit” OR “Im not fit” OR “I'm not fit” OR alcohol OR alcoholic OR #alcoholic OR “heavy drinker” OR “I am a drinker” OR “Im a drinker” OR “I'm a drinker” OR tobacco OR nicotine OR “I smoke” OR “used to smoke” OR “was a smoker” OR “am a smoker” OR “is a smoker” OR cigarettes OR “pack a day” OR “packs a day”
)
)
AND NOT

(kidney OR kidneys OR #kidney OR #kidneys OR CKD OR #CKD OR IgAN OR #IgAN OR iganephropathy OR #IgANephropathy OR #kidneydisease OR #chronickidneydisease OR #nephrology OR #kidneyhealth OR renal OR #renal OR #renaldisease OR ESRD OR #ESRD OR ESKD OR #ESKD OR #kidneyfailure OR #RenalArteryStenosis OR #kidneytransplant OR #dialysis OR #kidneyawareness OR #kidneydiseaseawareness OR #kidneywarrior OR adrenal OR #adrenal OR #adrenaltumor OR heart OR cardiac OR CHF OR #CHF OR “side effect” OR “side effects”)
)
)
OR
(// RENAL HYPERTENSION //
(“renal hypertension” OR #renalhypertension OR “hypertensive nephropathy” OR #hypertensivenephropathy OR “hypertensive kidney disease” OR #hypertensivekidneydisease OR “hypertension due to kidney” OR “high blood pressure due to kidney”)
OR
(
(“secondary hypertension” OR #secondaryhypertension OR #secondary #hypertension OR “high blood pressure” OR #highbloodpressure OR hypertension OR #hypertension OR “hypertensive crisis” OR #hypertensivecrisis OR “hypertensive emergency” OR #hypertensiveemergency OR #bloodpressure OR #highbloodpressure OR #hypertensiontreatment OR “antihypertensive drugs” OR antihypertensive OR #antihypertensive OR “DASH diet” OR #DASHdiet OR HBP OR #HBP OR #hypertensionawareness)
AND
(kidney OR kidneys OR #kidney OR #kidneys OR CKD OR #CKD OR IgAN OR #IgAN OR iganephropathy OR #IgANephropathy OR #kidneydisease OR #chronickidneydisease OR #nephrology OR #kidneyhealth OR renal OR #renal OR #renaldisease OR ESRD OR #ESRD OR ESKD OR #ESKD OR #kidneyfailure OR #RenalArteryStenosis OR #kidneytransplant OR #dialysis OR #kidneyawareness OR #kidneydiseaseawareness OR #kidneywarrior)
)
)
OR
(// CARDIAC HYPERTENSION //
(“cardiac hypertension” OR #cardiachypertension OR “hypertensive cardiac disease” OR #hypertensivecardiacdisease OR “hypertensive heart disease” OR #hypertensiveheartdisease)
OR
(
(“secondary hypertension” OR #secondaryhypertension OR #secondary #hypertension OR “high blood pressure” OR #highbloodpressure OR hypertension OR #hypertension OR “hypertensive crisis” OR #hypertensivecrisis OR “hypertensive emergency” OR #hypertensiveemergency OR #bloodpressure OR #highbloodpressure OR #hypertensiontreatment OR “antihypertensive drugs” OR antihypertensive OR #antihypertensive OR “DASH diet” OR #DASHdiet OR HBP OR #HBP OR #hypertensionawareness)
AND

(heart OR “heart disease” OR #heartdisease OR #heart OR “heart defects” OR #congenitalheartdisease OR atherosclerotic OR atherosclerosis OR ASCVD OR #ASCVD OR “heart failure” OR #heartfailure OR HF OR #HF OR HFpEF OR #HFpEF OR #HFrEF OR HFrEF OR cardiovascular OR “cv disease” OR “coronary artery disease” OR CAD OR #CAD OR cardiomyopathy OR #cardiomyopathy OR CHF OR #CHF OR arrhythmia OR #arrhythmia OR “atrial fibrillation” OR #afib OR afib OR #heartattack OR myocardial OR #MI OR MI OR angina OR #angina OR stroke OR #stroke OR ischemic OR “artery disease” OR #venousthromboembolism OR thromboembolism OR “deep vein thrombosis” OR #DVT OR DVT OR embolism OR #pulmonaryembolism OR #heartvalvedisease OR “mitral valve” OR aortic OR #aorticaneurysm OR #cardiacarrest OR “sudden cardiac death” OR #suddencardiacdeath OR #vasculardisease OR vascular OR “endothelial dysfunction” OR #endothelialdysfunction OR “blood clot” OR #bloodclot OR “lipid disorders” OR Hypercholesterolemia OR #Hypercholesterolemia OR hypercholesterolaemia OR #hypercholesterolaemia OR “high cholesterol” OR triglycerides OR #cardiacbiomarkers OR troponin OR “brain natriuretic peptide” OR “c-reactive protein” OR homocysteine OR electrocardiogram OR ECG OR EKG OR echocardiography OR “cardiac stress test” OR angiography OR “coronary angioplasty” OR “stent placement” OR “cardiac bypass surgery” OR pacemaker OR “implantable cardioverter defibrillator” OR arteriosclerosis OR #arteriosclerosis OR #congestiveheartdisease OR chd OR #CHD OR “narrowing coronary arteries” OR “blockage of coronary arteries”)
)
)
)
AND
(// PATIENT EXPERIENCE TERMS //
(“I have hypertension” OR “I have” OR “my blood pressure” OR “my BP” OR “my B.P.” OR “my B.P” OR “my high” OR “my blood” OR “my HBP” OR “my hypertension” OR “I was” OR “I am” OR “I struggle with” OR “my experiences with hypertension” OR “I am living with” OR “Im living with” OR “I was diagnosed” OR “Im diagnosed” OR “I'm diagnosed” OR “my dx” OR “my diagnosis” OR #livingwithhypertension OR #livingwithhighbloodpressure OR “I have high blood pressure” OR “my experiences withhigh blood pressure” OR “diagnosed me with high blood pressure” OR “my high blood pressure” OR “I have high BP” OR “my experiences with high BP” OR “I was diagnosed with high BP” OR “diagnosed me” OR “my high BP” OR “I have resistant” OR “my experiences with” OR “my resistant hypertension” OR “my headaches” OR “my chestpain” OR “my breathing” OR “my urine” OR “my heartbeat” OR “my results” OR “my labs” OR “my lab results” OR “my test results” OR “my tests came back” OR “my levels” OR “my pressure”) OR
// SPECIALISTS //
(“my cardiologist” OR “my endocrinologist” OR “my kidney doctor” OR “my specialist” OR “my nephrologist” OR “my PCP” OR “my doctor” OR “my neph” OR “my primary care” OR “my GP” OR “my doc” OR “my DR” OR “my physician” OR “my NP” OR “my nurse” OR “my pharmacist” OR “my pharmacy” OR “my doctors” OR “went to doctor” OR “went to physician” OR “went to doctors” OR “going to doctor” OR “going to the doctor” OR “going to the doctors”) OR
// STANDARD - DISEASE //
(“my disease” OR “my illness” OR “my condition” OR “I have a disease” OR “I have a condition” OR “i have an illness” OR “I have a diagnosis” OR “I am diagnosed” OR “I'm

diagnosed” OR “I got diagnosed” OR “i have been diagnosed” OR “I have severe” OR “I have moderate” OR “I have mild” OR “I have stage” OR “my symptom” OR “my first symptom” OR “my symptoms” OR “my first symptoms” OR “my pain” OR “my relapse” OR “I relapsed” OR “I have relapsed” OR “I am relapsing” OR “I am worsening” OR “my flare-up” OR “I flared up” OR “I flared” OR “I am flaring” OR “my life with” OR “my struggles with” OR “I'm living with” OR “my battle with” OR “my fight with” OR “i live with” OR “how i dealt with” OR “how i deal with” OR “I am a patient”) OR
// STANDARD - TREATMENT //
(“my drug” OR “my pill” OR “my med” OR “my medication” OR “my medicine” OR “my meds” OR “my prescription” OR “my rx” OR “my injection” OR “my infusion” OR “my shot” OR “my treatment” OR “my surgery” OR “i started on” OR “I was put on” OR “my dosage” OR “my dose” OR “my diet”) OR
// STANDARD - DOCTORS & APPOINTMENTS //
(“doctor suggested” OR “dr suggest” OR “dr recommend” OR “doctor told me” OR “doc told me” OR “physican told me” OR “nurse told me” OR “specialist told me” OR “my appointment” OR “I had an appointment” OR “my dr appointment” OR “my specialist appointment” OR “my dr appt” OR “my specialist appt” OR “I saw a” OR “I went to a” OR “my insurance” OR “my hospital bill” OR “my copay” OR “my out-of-pocket” OR “my co-pay”)
)
)
AND
(geo:”United States” OR geo:”United Kingdom” OR geo:Canada OR geo:Germany OR geo:Australia OR geo:France OR geo:Spain OR geo:Italy OR geo:Japan OR geo:Brazil OR geo:”South Korea”)

**Table S1**. Comorbidity area mention volumes across total core sample (n=81,368)[a]

| Comorbidity | Total (%) |
|---|---|
| Cardiovascular conditions | 24 |
| Metabolic disorders | 19 |
| Psychological conditions | 9 |
| Renal disorders | 7 |
| Respiratory diseases | 4 |
| Pregnancy issues | 3 |
| Cancers | 3 |
| Musculosceletal disorders | 2 |
| Ophthalmologic conditions | 1 |
| Thyroid conditions | 1 |
| Autoimmune conditions | <1 |
| Hepatological conditions | <1 |
| Gastrointestinal conditions | <1 |
| Dermatologic conditions | <1 |

[a]Core samples excludes China due to country data transfer regulations. Sample shown to nearest '00.

**Table S2**. Total mentions of comorbidities in European area mention volumes across total core sample (n=81,368)

| Country | n | Comorbidities n (%) |
|---|---|---|
| Europe | 16,528 | 9,588 (56%) |
| Spain | 5,059 | 3,694 (73%) |
| France | 2,686 | 1,704 (63%) |
| UK | 4,589 | 2,293 (50%) |
| Germany | 4,194 | 1,897 (45%) |
| Japan | 5,486 | 2,555 (47%) |
| China | 5,000 | 3,064 (61%) |
| South Korea | 3,517 | 2,189 (62%) |

| | | |
|---|---|---|
| Australia | 915 | 459 (50%) |
| North America | 38,737 | 16,922 (44%) |
| United States | 36,307 | 15,681 (43%) |
| Canada | 2,430 | 1,241 (51%) |
| Brazil | 11,209 | 4,258 (38%) |

**Table S3**. Comorbidity area mention volumes by age group across core sample (n=8,096)[a]

| Life impact area mentions | <40 years old (%) | 40+ years old (%) |
|---|---|---|
| Cardiovascular conditions | 28 | 34 |
| Diabetes (metabolic disorder) | 16 | 26 |
| Obesity (metabolic disorder) | 17 | 12 |
| Psychological conditions | 11 | 9 |

[a]Core samples excludes China due to country data transfer regulations. Sample shown to nearest ’00.

**Table S4**. Individual symptom mentions volumes (of >1%) across total core sample (n=81,368)[a]

| Symptom concepts | Total Matches (%) |
|---|---|
| Joint & muscle pain | 5 |
| Headaches & migraines | 4 |
| Hearing changes & loss | 4 |
| Vision changes & loss | 3 |
| Fatigue | 2 |
| Dizziness | 2 |
| Edema & swelling | 2 |
| Sweating & overheating | 2 |
| Breathing difficulties | 2 |
| Insomnia | 1 |
| Sleep apnea | 1 |
| Brain fog & confusion | 1 |
| Skin rashes | 1 |
| Chills | 1 |

| | |
|---|---|
| Digestive issues | 1 |

[a]Core samples excludes China due to country data transfer regulations. Sample shown to nearest '00.

**Table S5**. Symptom area mentions by controlled vs uncontrolled/ resistant patients across core sample (n=18,705)[a]

| Symptom Concepts | Uncontrolled or Resistant Individuals (%) | Controlled Individuals (%) |
|---|---|---|
| Aches & pains | 16 | 10 |
| Sensory disturbances | 12 | 9 |
| Fatigue & sleep issues | 11 | 6 |
| Dizziness & fogginess | 7 | 4 |
| Poor thermoregulation | 4 | 4 |
| Rashes, swelling & bleeding | 5 | 3 |
| Breathing issues | 4 | 3 |

[a]Core samples excludes China due to country data transfer regulations. Sample shown to nearest '00.

**Table S6**. QoL impact area mention volumes by gender across core sample (Total n=22,394; Male n=10,875; Female n=11,519)[a]

| Life impact area mentions | Male (%) | Female (%) | Individual life impacts mentions | Male (%) | Female (%) |
|---|---|---|---|---|---|
| Everyday Life | 17 | 21 | Time spent on medical appointments | 6 | 9 |
| | | | Sleep disruption | 6 | 6 |
| | | | Time spent researching online | 5 | 3 |
| | | | Functional limitations | 3 | 2 |
| | | | Household task difficulties | 2 | 2 |
| Emotional Balance | 17 | 20 | Worry and Anxiety | 9 | 11 |
| | | | Sadness and depression | 4 | 5 |
| | | | Fear | 3 | 4 |

| | | | | | |
|---|---|---|---|---|---|
| | | | Shock and disbelief | 2 | 1 |
| Work/ Education | 10 | 8 | Impact on job performance | 9 | 8 |
| Social Connections | 6 | 5 | Impact on family & friendships | 3 | 3 |
| | | | Ability to maintain a social life | 2 | 1 |
| Financial Health | 6 | 5 | Expense cost concerns | 3 | 2 |
| | | | Insurance coverage | 2 | 2 |

[a]Core samples excludes China due to country data transfer regulations. Sample shown to nearest ’00.

**Table S7**. QoL impact mentions by focus comorbidities across core sample (n=39,171)[a]

| Life impact area mentions | Primary hypertension (%) | Hypertension with cardiovascular comorbidities (%) | Hypertension with renal comorbidities (%) |
|---|---|---|---|
| Everyday Life | 23 | 28 | 26 |
| Emotional Balance | 22 | 23 | 22 |
| Work/ Education | 12 | 12 | 12 |
| Social Connections | 7 | 8 | 7 |
| Financial Health | 7 | 7 | 6 |

[a]Core samples excludes China due to country data transfer regulations. Sample shown to nearest ’00.

**Figure S1**. QoL impact mentions by ethnicity/ race across US sample: Qualitative overview (n=794)[a]

| | Black | White | Hispanic | Asian |
|---|---|---|---|---|
| **Everyday Life** | • Time spent on online research<br>• Disrupted sleep | • Continuous fatigue<br>• Regular clinic/ ER visits | • Emergency medical appt.<br>• Regular specialist visits | • Disrupted sleep<br>• Increased daytime fatigue<br>• Reduced productivity/ focus |
| **Emotional Balance** | • Anxiety & worry<br>• Specific fears in pregnancy<br>• Frustration from HCP dismissal | • Anxiety & worry<br>• Confusion | • Anxiety & worry<br>• Resignation around health decline | • Anxiety & worry<br>• Shame & guilt of lifestyle 'causes' |
| **Work/ Education** | • Concern over work life impacting health<br>• Low energy & motivation | | | • Fatigue impacting work<br>• Fear of work & school-related stress impacting health |
| **Social Connections** | | • Emotional toll on caregivers | | |
| **Finances** | | | | |

| Cell color key | Higher proportion of posts mentioning impact area | Mid proportion of posts mentioning impact area | Lower proportion of posts mentioning impact area |
|---|---|---|---|

[a]Sample: United States posts with ethnicity information. Sample shown to nearest '00.
